# Neighborhood and Environmental Factors and Post-Discharge Healthcare Utilization in HFpEF: A Retrospective Cohort Study

**DOI:** 10.64898/2026.07.15.26358204

**Authors:** Yeabsra Aleligne, Erick Romero, Clodomir Santana, Julie T. Bidwell, Javier E. López, Miriam Nuño, Imo Ebong, Leighton Izu, David A. Liem, Nipavan Chiamvimonvat, Martin Cadeiras

## Abstract

**Background:** Neighborhood-level social determinants of health influence cardiovascular outcomes; however, their association with post-discharge healthcare utilization in heart failure with preserved ejection fraction (HFpEF) remains incompletely defined.

**Methods:** We conducted a retrospective cohort study of 6,702 adults hospitalized for HFpEF (2014 to 2022). Patients were assigned to one of the four neighborhood environments (NEnv-1 to NEnv-4) using a validated clustering framework based on ZIP code–level socioeconomic variables. The primary outcome was time to first HF readmission, evaluated within prespecified post-discharge intervals (0–30 days, >30–90 days, and >90–365 days). The secondary outcomes included HF-related healthcare re-encounters and HF hospitalization burden (0, 1, or ≥2 admissions). Cox proportional hazard and multinomial logistic regression models were used.

**Results:** Neighborhood environment was independently associated with post-discharge outcomes with distinct temporal patterns. Early (0–30 days) HF readmission risk was higher in NEnv-3 (aHR, 1.63) and NEnv-4 (aHR, 1.76), with similar increases in HF-related re-encounters (aHR, 1.72 and 1.84) persisting through the >30–90-day interval. In contrast, NEnv-2 demonstrated a delayed-risk pattern, with the highest risk occurring in the >90–365-day interval (readmission aHR, 3.42; re-encounter aHR, 3.45). All non-reference environments were associated with a higher likelihood of at least one post-index HF admission (aOR range, 1.84–2.24). NEnv-4 uniquely demonstrated a higher odds of recurrent hospitalization (≥2 vs. 1 admission; aOR, 1.64).

**Conclusions:** Neighborhood environment is associated with distinct, time-dependent patterns of HF utilization in HFpEF, including early, delayed, and recurrent risks. Incorporating neighborhood context may help identify when patients with HFpEF are most vulnerable after discharge and guide the timing of post-discharge interventions.

## Introduction

Heart failure (HF) affects more than 6 million adults in the United States and remains a major contributor to morbidity, mortality, and healthcare utilization.^1,2^ In particular, heart failure with preserved ejection fraction (HFpEF) now accounts for more than half of HF cases^3^ and continues to increase in prevalence, driven by aging populations and cardiometabolic comorbidity.^4^ HFpEF is associated with high post-discharge healthcare utilization, with approximately 20% of patients readmitted within 30 days,^4,5^ and more than 50% within 1 year.^4,5^ Despite this burden, determinants of post-discharge outcomes during the vulnerable phase^6^ remain incompletely defined.^7–9^

HFpEF is increasingly recognized as a heterogeneous syndrome composed of distinct clinical phenotypes that differ in comorbidity burden, systemic inflammation, cardiac structure, prognosis, and treatment response.^10–13^ Data-driven phenomapping studies have improved the characterization of HFpEF subgroups; however, these approaches largely define phenotypes using downstream clinical and biological features.^10,12,13^ Little is known about whether upstream social and environmental exposures shape HFpEF phenotype expression and contribute to differences in post-discharge healthcare utilization.

Social determinants of health, which are the conditions in which individuals are born, live, and work, are key contributors to cardiovascular disease risk and outcomes.^14^ Adverse social conditions, including poverty and limited access to care, impair HF self-management and care delivery.^7,9^ These factors are associated with reduced medication adherence and barriers to longitudinal care,^2,7^ as well as increased hospitalization and mortality risk.^2,15,16^ Rurality-related barriers further contribute to disparities in outcomes.^17^

Prior studies have largely focused on individual factors, such as neighborhood deprivation, income,^15,16^ or rurality.^17^ This approach may obscure how co-occurring social, demographic, economic, healthcare access, and built-environment exposures jointly influence HFpEF phenotypes and downstream outcomes. A multidimensional characterization of neighborhood environments may better capture these interacting systems-level exposures.^8^

Previously, we developed a clustering framework to classify ZIP Code Tabulation Areas into four multidimensional neighborhood environments (NEnv-1 through NEnv-4) using socioeconomic indicators across major domains of social determinants of health.^18,19^ These environments capture co-occurring social and built-environment exposures and have been associated with distinct HFpEF phenotypes and mortality risk.^20^ However, whether these neighborhood environment phenotypes are associated with post-discharge healthcare utilization, including HF readmissions, healthcare re-encounters, and cumulative hospitalization burden, remains unknown.

In this study, we evaluated the association between neighborhood environment phenotypes and post-discharge outcomes in a large HFpEF cohort. Specifically, we assessed HF readmissions, healthcare re-encounters, and cumulative hospitalization burden across prespecified post-discharge intervals (0–30, >30–90, and >90–365 days). We hypothesized that neighborhood environment would be associated with differential, time-dependent patterns of post-discharge healthcare utilization.

## Methods

### Study Design and Population

This retrospective cohort study included adults hospitalized for HFpEF at the University of California Davis Medical Center between January 2014 and December 2022. The study was approved by the University of California, Davis Institutional Review Board, and the requirement for informed consent was waived due to the retrospective, de-identified nature of the data.

HFpEF was defined as LVEF ≥50% with an elevated BNP level of ≥100 pg/mL and corresponding ICD-9 or ICD-10 codes. Exclusion criteria included LVEF <50%, prior heart transplantation or use of a left ventricular assist device, pulmonary arterial hypertension therapy, and missing ZIP codes or hospitalization data. The final cohort included 6,702 patients (Figure 1). Baseline demographic, clinical, laboratory, and medication data were obtained from electronic health records. Index hospitalization was defined as the first HF hospitalization during the study period. Patients were eligible for post-discharge outcome analyses only after discharge from the index HF hospitalization.

**Figure 1.**
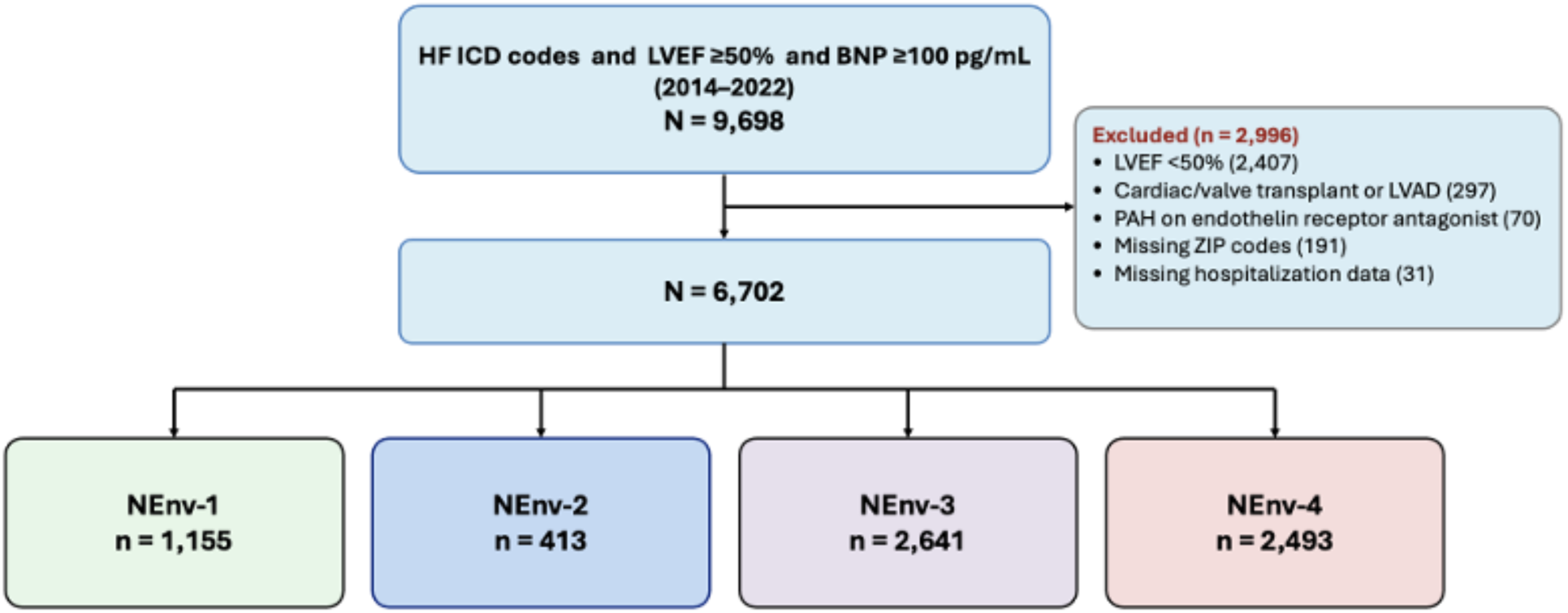
Study Cohort Flow Diagram. Flow diagram of cohort derivation from 9,698 patients identified with heart failure with preserved ejection fraction. Exclusions included left ventricular ejection fraction <50%, prior heart transplantation or left ventricular assist device, pulmonary arterial hypertension therapy, missing ZIP code, or missing hospitalization data. The final analytic cohort included 6,702 patients, who were assigned to one of four neighborhood environments (NEnv-1 to NEnv-4) based on residential ZIP code.

### Neighborhood Environment Assignment

Neighborhood environment was defined using a previously developed and validated ZIP Code Tabulation Area (ZCTA)-level clustering framework.^18–20^ Briefly, ZCTA-level variables from the American Community Survey were selected to capture multiple domains of social determinants of health, including economic stability, education, healthcare access, and neighborhood and built environment. These variables included measures of poverty, employment, median household income, internet access, health insurance coverage, educational attainment, land area, average family size, dependency ratios, sex distribution, primary language, race and ethnicity, and the proportion of the foreign-born population.

As previously described,^18–20^ an unsupervised machine learning approach (k-means clustering) was applied to classify ZCTAs into four neighborhood environments (NEnv-1 to NEnv-4).

Patients were assigned to neighborhood environments by linking residential ZIP codes to corresponding ZCTA-level cluster assignments. These environments represent multidimensional profiles of social and built-environment characteristics rather than single-domain measures.^20^

### Outcomes

The primary outcome was time to first HF readmission after discharge, analyzed within prespecified post-discharge intervals (0–30 days, >30–90 days, and >90–365 days). Secondary outcomes included HF-related healthcare re-encounters and HF hospitalization burden. HF readmission was defined as a subsequent HF hospitalization occurring more than 24 h after discharge from the index hospitalization. HF-related healthcare re-encounters were defined as a composite of HF-related emergency department visits, observation stays, or inpatient readmissions. The HF hospitalization burden after the index HF hospitalization was categorized as 0, 1, or ≥2 subsequent HF admissions.

### Statistical Analysis

Continuous variables are presented as mean ± SD or median [interquartile range] based on distribution, and categorical variables as counts and percentages. Between-group comparisons were performed using one-way analysis of variance or the Kruskal–Wallis test for continuous variables and Pearson χ² or Fisher’s exact test for categorical variables, as appropriate. All tests were 2-sided, with P<0.05 considered statistically significant.

Time-to-event outcomes, including HF readmission and HF-related healthcare re-encounters, were analyzed using Cox proportional hazards models. The proportional hazards assumption, assessed by log-minus-log plots, was violated. Follow-up was therefore divided into prespecified, non-overlapping post-discharge intervals (0–30, >30–90, and >90–365 days), with separate Cox models fitted within each interval among patients who were event-free at the start of that interval. Hazard ratios represent interval-specific risks conditional on remaining event-free at the start of each interval. Patients entered the risk set at the start of each interval and were censored at the event or at the end of the time period. Adjusted hazard ratios and odds ratios are reported with 95% confidence intervals.

Multivariable models were adjusted for clinically relevant covariates selected a priori, including age, sex, race/ethnicity, hypertension, diabetes mellitus, hyperlipidemia, chronic kidney disease, atrial fibrillation/flutter, chronic obstructive pulmonary disease, ischemic heart disease, hypothyroidism, body mass index, log-transformed BNP, systolic blood pressure, heart rate, and left ventricular ejection fraction. Race/ethnicity was included as a categorical covariate in adjusted models and was consolidated into White, Black, Asian, Latinx/Hispanic, and Other/Unknown categories to reduce sparse-cell instability. To minimize multicollinearity, correlated variables were not entered simultaneously; body mass index was used instead of obesity, chronic kidney disease instead of eGFR or creatinine, and systolic blood pressure instead of multiple blood pressure measures. NEnv-1 was used as a reference group.

Complete case analysis was performed without imputation. Covariates with substantial missingness, including QTc interval and medication variables, were excluded from the adjusted models to preserve the sample size and reduce potential selection bias. Medication variables were excluded due to substantial missingness and potential confounding by indication.

The HF hospitalization burden after the index hospitalization was analyzed using multinomial logistic regression. Outcomes were modeled as 1 versus 0 HF admissions and ≥2 versus 1 HF admissions. The models were adjusted for the same clinical covariates and follow-up duration. Adjusted hazard ratios and adjusted odds ratios were reported with 95% confidence intervals.

All analyses were performed using IBM SPSS Statistics for Mac, version 30.0 (IBM Corp., Armonk, NY).

## Results

### Study Cohort and Baseline Characteristics

A total of 6,702 patients were included in the analytical cohort, with 1,155 (17.2%), 413 (6.2%), 2,641 (39.4%), and 2,493 (37.2%) patients in the NEnv-1 through NEnv-4 groups, respectively. The baseline characteristics varied significantly across neighborhood environments (Table 1), with distinct demographic and clinical phenotypes.

**Table 1.**
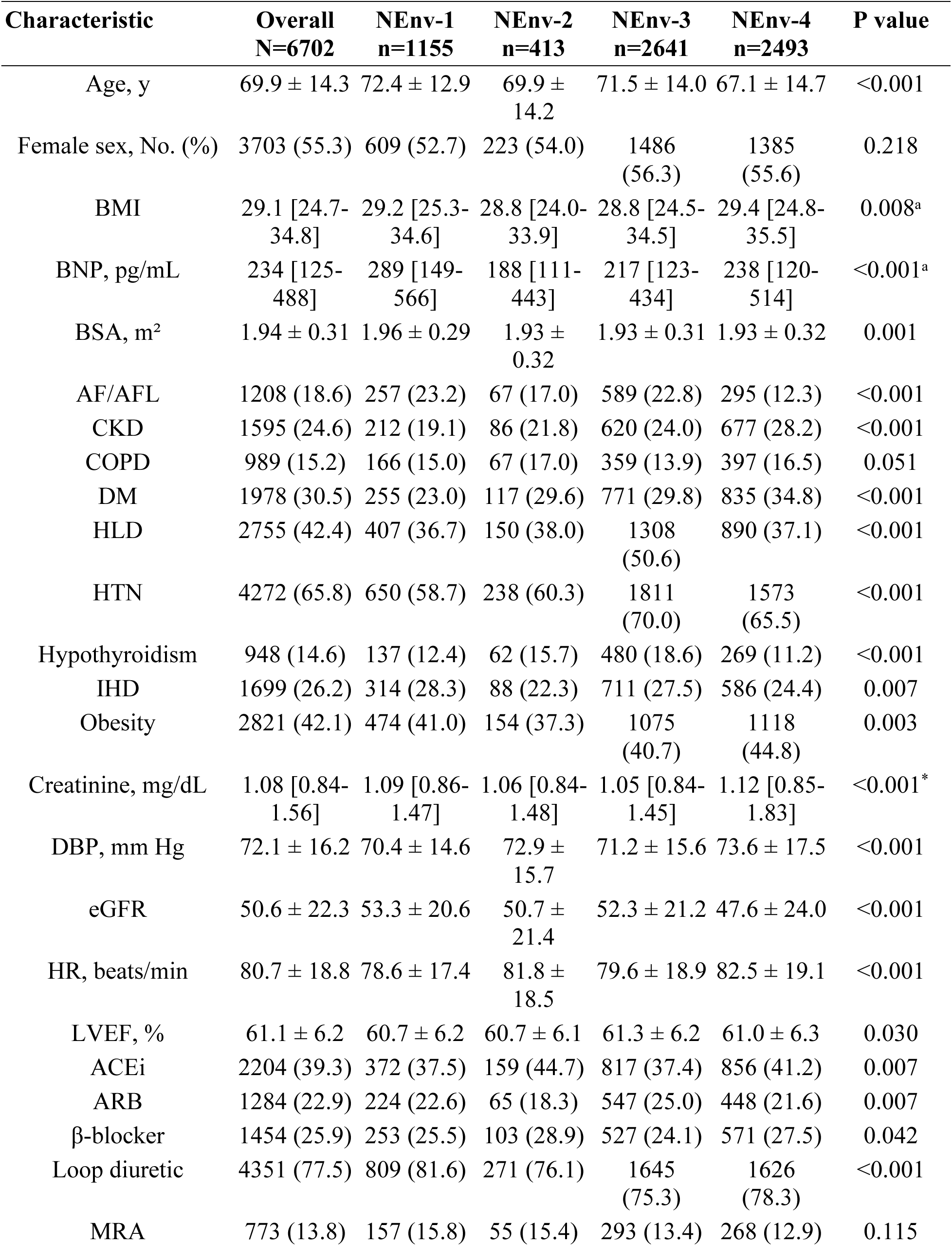
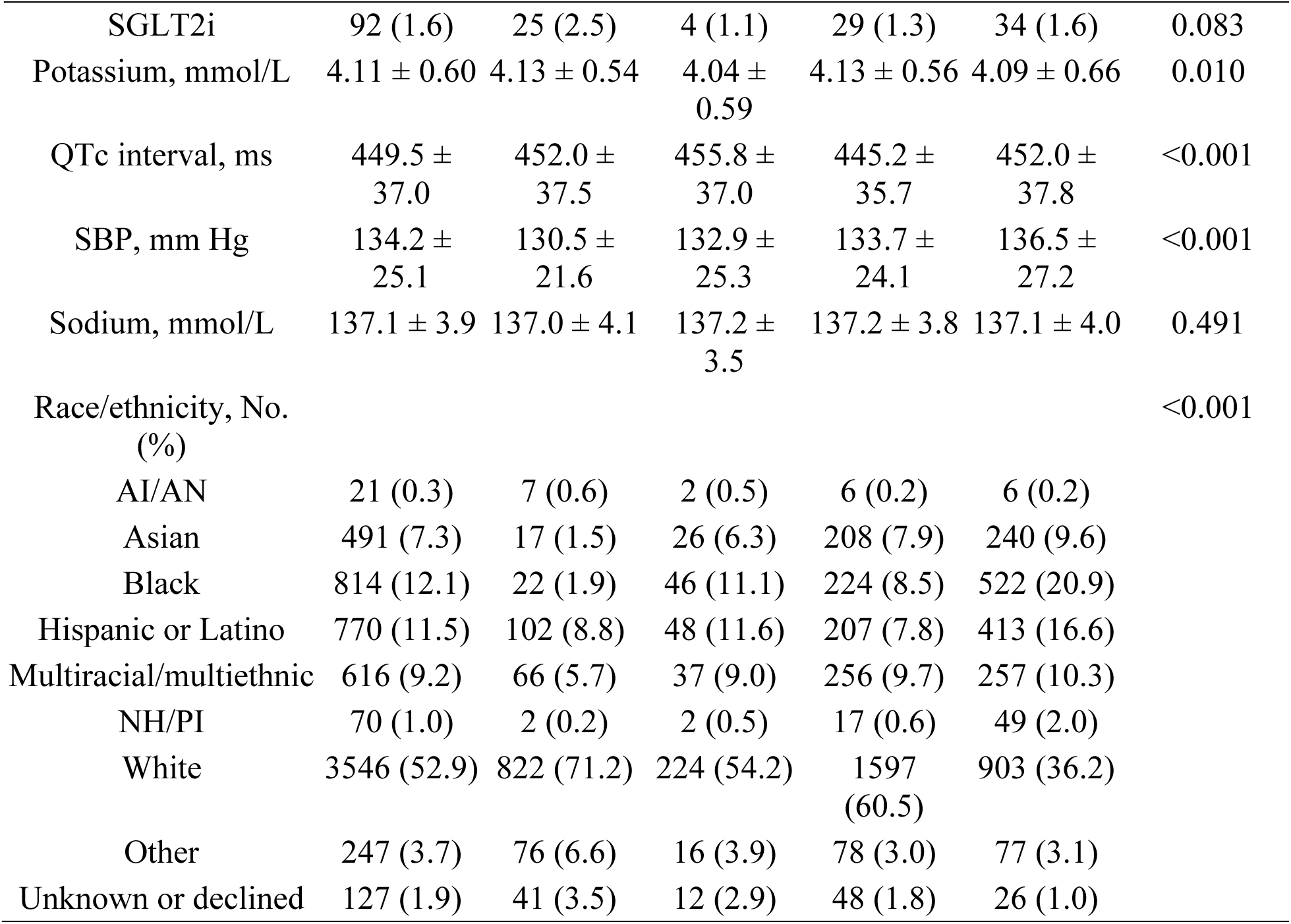
Baseline Characteristics by Neighborhood Environment. Baseline demographic, clinical, laboratory, and medication characteristics of the study cohort (N=6,702), stratified by neighborhood environment (NEnv-1 to NEnv-4). Continuous variables are presented as mean ± standard deviation or median [interquartile range], and categorical variables as number (%). Percentages are column-based and calculated using non-missing denominators. P values reflect comparisons across groups using one-way analysis of variance or Kruskal–Wallis tests for continuous variables, and χ² or Fisher exact tests for categorical variables, as appropriate. **Abbreviations:** ACEi, angiotensin-converting enzyme inhibitor; AF/AFL, atrial fibrillation or flutter; AI/AN, American Indian or Alaska Native; ARB, angiotensin receptor blocker; BMI, body mass index; BNP, brain natriuretic peptide; BSA, body surface area; CKD, chronic kidney disease; COPD, chronic obstructive pulmonary disease; DBP, diastolic blood pressure; DM, diabetes mellitus; eGFR, estimated glomerular filtration rate (mL/min/1.73 m²); HLD, hyperlipidemia; HR, heart rate; HTN, hypertension; IHD, ischemic heart disease; IQR, interquartile range; LVEF, left ventricular ejection fraction; MRA, mineralocorticoid receptor antagonist; NEnv, neighborhood environment; NH/PI, Native Hawaiian or other Pacific Islander; QTc, corrected QT interval; SBP, systolic blood pressure; SGLT2i, sodium-glucose cotransporter 2 inhibitor.

| **Characteristic** | **Overall**<br>**N=6702** | **NEnv-1**<br>**n=1155** | **NEnv-2**<br>**n=413** | **NEnv-3**<br>**n=2641** | **NEnv-4**<br>**n=2493** | **P value** |
| --- | --- | --- | --- | --- | --- | --- |
| Age, y | 69.9 ± 14.3 | 72.4 ± 12.9 | 69.9 ± 14.2 | 71.5 ± 14.0 | 67.1 ± 14.7 | <0.001 |
| Female sex, No. (%) | 3703 (55.3) | 609 (52.7) | 223 (54.0) | 1486 (56.3) | 1385 (55.6) | 0.218 |
| BMI | 29.1 [24.7-34.8] | 29.2 [25.3-34.6] | 28.8 [24.0-33.9] | 28.8 [24.5-34.5] | 29.4 [24.8-35.5] | 0.008<sup>a</sup> |
| BNP, pg/mL | 234 [125-488] | 289 [149-566] | 188 [111-443] | 217 [123-434] | 238 [120-514] | <0.001<sup>a</sup> |
| BSA, m<sup>2</sup> | 1.94 ± 0.31 | 1.96 ± 0.29 | 1.93 ± 0.32 | 1.93 ± 0.31 | 1.93 ± 0.32 | 0.001 |
| AF/AFL | 1208 (18.6) | 257 (23.2) | 67 (17.0) | 589 (22.8) | 295 (12.3) | <0.001 |
| CKD | 1595 (24.6) | 212 (19.1) | 86 (21.8) | 620 (24.0) | 677 (28.2) | <0.001 |
| COPD | 989 (15.2) | 166 (15.0) | 67 (17.0) | 359 (13.9) | 397 (16.5) | 0.051 |
| DM | 1978 (30.5) | 255 (23.0) | 117 (29.6) | 771 (29.8) | 835 (34.8) | <0.001 |
| HLD | 2755 (42.4) | 407 (36.7) | 150 (38.0) | 1308 (50.6) | 890 (37.1) | <0.001 |
| HTN | 4272 (65.8) | 650 (58.7) | 238 (60.3) | 1811 (70.0) | 1573 (65.5) | <0.001 |
| Hypothyroidism | 948 (14.6) | 137 (12.4) | 62 (15.7) | 480 (18.6) | 269 (11.2) | <0.001 |
| IHD | 1699 (26.2) | 314 (28.3) | 88 (22.3) | 711 (27.5) | 586 (24.4) | 0.007 |
| Obesity | 2821 (42.1) | 474 (41.0) | 154 (37.3) | 1075 (40.7) | 1118 (44.8) | 0.003 |
| Creatinine, mg/dL | 1.08 [0.84-1.56] | 1.09 [0.86-1.47] | 1.06 [0.84-1.48] | 1.05 [0.84-1.45] | 1.12 [0.85-1.83] | <0.001<sup>*</sup> |
| DBP, mm Hg | 72.1 ± 16.2 | 70.4 ± 14.6 | 72.9 ± 15.7 | 71.2 ± 15.6 | 73.6 ± 17.5 | <0.001 |
| eGFR | 50.6 ± 22.3 | 53.3 ± 20.6 | 50.7 ± 21.4 | 52.3 ± 21.2 | 47.6 ± 24.0 | <0.001 |
| HR, beats/min | 80.7 ± 18.8 | 78.6 ± 17.4 | 81.8 ± 18.5 | 79.6 ± 18.9 | 82.5 ± 19.1 | <0.001 |
| LVEF, % | 61.1 ± 6.2 | 60.7 ± 6.2 | 60.7 ± 6.1 | 61.3 ± 6.2 | 61.0 ± 6.3 | 0.030 |
| ACEi | 2204 (39.3) | 372 (37.5) | 159 (44.7) | 817 (37.4) | 856 (41.2) | 0.007 |
| ARB | 1284 (22.9) | 224 (22.6) | 65 (18.3) | 547 (25.0) | 448 (21.6) | 0.007 |
| β-blocker | 1454 (25.9) | 253 (25.5) | 103 (28.9) | 527 (24.1) | 571 (27.5) | 0.042 |
| Loop diuretic | 4351 (77.5) | 809 (81.6) | 271 (76.1) | 1645 (75.3) | 1626 (78.3) | <0.001 |
| MRA | 773 (13.8) | 157 (15.8) | 55 (15.4) | 293 (13.4) | 268 (12.9) | 0.115 |
| SGLT2i | 92 (1.6) | 25 (2.5) | 4 (1.1) | 29 (1.3) | 34 (1.6) | 0.083 |
| Potassium, mmol/L | 4.11 ± 0.60 | 4.13 ± 0.54 | 4.04 ± 0.59 | 4.13 ± 0.56 | 4.09 ± 0.66 | 0.010 |
| QTc interval, ms | 449.5 ± 37.0 | 452.0 ± 37.5 | 455.8 ± 37.0 | 445.2 ± 35.7 | 452.0 ± 37.8 | <0.001 |
| SBP, mm Hg | 134.2 ± 25.1 | 130.5 ± 21.6 | 132.9 ± 25.3 | 133.7 ± 24.1 | 136.5 ± 27.2 | <0.001 |
| Sodium, mmol/L | 137.1 ± 3.9 | 137.0 ± 4.1 | 137.2 ± 3.5 | 137.2 ± 3.8 | 137.1 ± 4.0 | 0.491 |
| Race/ethnicity, No. (%) |  |  |  |  |  | <0.001 |
| AI/AN | 21 (0.3) | 7 (0.6) | 2 (0.5) | 6 (0.2) | 6 (0.2) |  |
| Asian | 491 (7.3) | 17 (1.5) | 26 (6.3) | 208 (7.9) | 240 (9.6) |  |
| Black | 814 (12.1) | 22 (1.9) | 46 (11.1) | 224 (8.5) | 522 (20.9) |  |
| Hispanic or Latino | 770 (11.5) | 102 (8.8) | 48 (11.6) | 207 (7.8) | 413 (16.6) |  |
| Multiracial/multiethnic | 616 (9.2) | 66 (5.7) | 37 (9.0) | 256 (9.7) | 257 (10.3) |  |
| NH/PI | 70 (1.0) | 2 (0.2) | 2 (0.5) | 17 (0.6) | 49 (2.0) |  |
| White | 3546 (52.9) | 822 (71.2) | 224 (54.2) | 1597 (60.5) | 903 (36.2) |  |
| Other | 247 (3.7) | 76 (6.6) | 16 (3.9) | 78 (3.0) | 77 (3.1) |  |
| Unknown or declined | 127 (1.9) | 41 (3.5) | 12 (2.9) | 48 (1.8) | 26 (1.0) |  |

NEnv-1 was characterized by an older population (mean age 72.4±12.9 years; P<0.001) and a predominance of White patients (71.2%; P<0.001). This group had the highest prevalence of atrial fibrillation/flutter (23.2%) and ischemic heart disease (28.3%), along with a lower prevalence of diabetes mellitus (23.0%) and chronic kidney disease (19.1%) relative to other environments.

The NEnv-2 had intermediate demographic and clinical features. The mean age was 69.9±14.2 years, with a more diverse racial and ethnic composition (54.2% White, 11.1% Black, 11.6% Hispanic or Latino). The comorbidity burden was moderate across all conditions, including diabetes mellitus (29.6%), chronic kidney disease (21.8%), atrial fibrillation/flutter (17.0%), hypertension (60.3%), and hyperlipidemia (38.0%).

NEnv-3 showed a comorbidity profile that was dominated by cardiometabolic conditions. This group had the highest prevalence of hypertension (70.0%) and hyperlipidemia (50.6%) (both P < 0.001), along with a relatively high prevalence of atrial fibrillation/flutter (22.8%). Demographic features were similar to those of NEnv-1, with an older population (mean age 71.5 ± 14.0 years) and a majority White population (60.5%).

NEnv-4 represented the youngest and most racially and ethnically diverse population (mean age 67.1±14.7 years; P<0.001), with higher proportions of Black (20.9%), Hispanic or Latino (16.6%), Asian (9.6%), and Native Hawaiian or Other Pacific Islander patients (2.0%) (P<0.001). This environment had the highest prevalence of diabetes mellitus (34.8%), chronic kidney disease (28.2%), and obesity (44.8%), whereas atrial fibrillation/flutter was the least prevalent (12.3%; P<0.001). The prevalence of COPD did not differ significantly across the groups (P=0.051), whereas that of ischemic heart disease varied modestly (P=0.007).

Laboratory and physiological measures differed across neighborhood environments (Table 1). NEnv-1 had the highest median BNP levels (289 [149–566] pg/mL; P<0.001), whereas NEnv-4 had the lowest mean eGFR (47.6±24.0 mL/min/1.73 m²; P<0.001) and the highest systolic blood pressure (136.5±27.2 mm Hg; P<0.001). Differences in BMI (P=0.008) and LVEF (P=0.030) were modest across the groups.

Medication use varied modestly across neighborhood environments, including ACEi, ARB, β-blockers, and loop diuretics (all P<0.05), whereas MRA and SGLT2i use was low and similar across the groups.

### Heart Failure Readmission

Neighborhood environment was significantly associated with HF readmission across all post-discharge intervals (Figure 2A–C; Table 2). Within the first 30 days after discharge, the adjusted risk of readmission was higher in NEnv-3 (aHR, 1.63; 95% CI, 1.12–2.37; P=0.011) and NEnv-4 (aHR, 1.76; 95% CI, 1.21–2.57; P=0.003), whereas NEnv-2 was not associated with an increased early risk.

**Figure 2.**
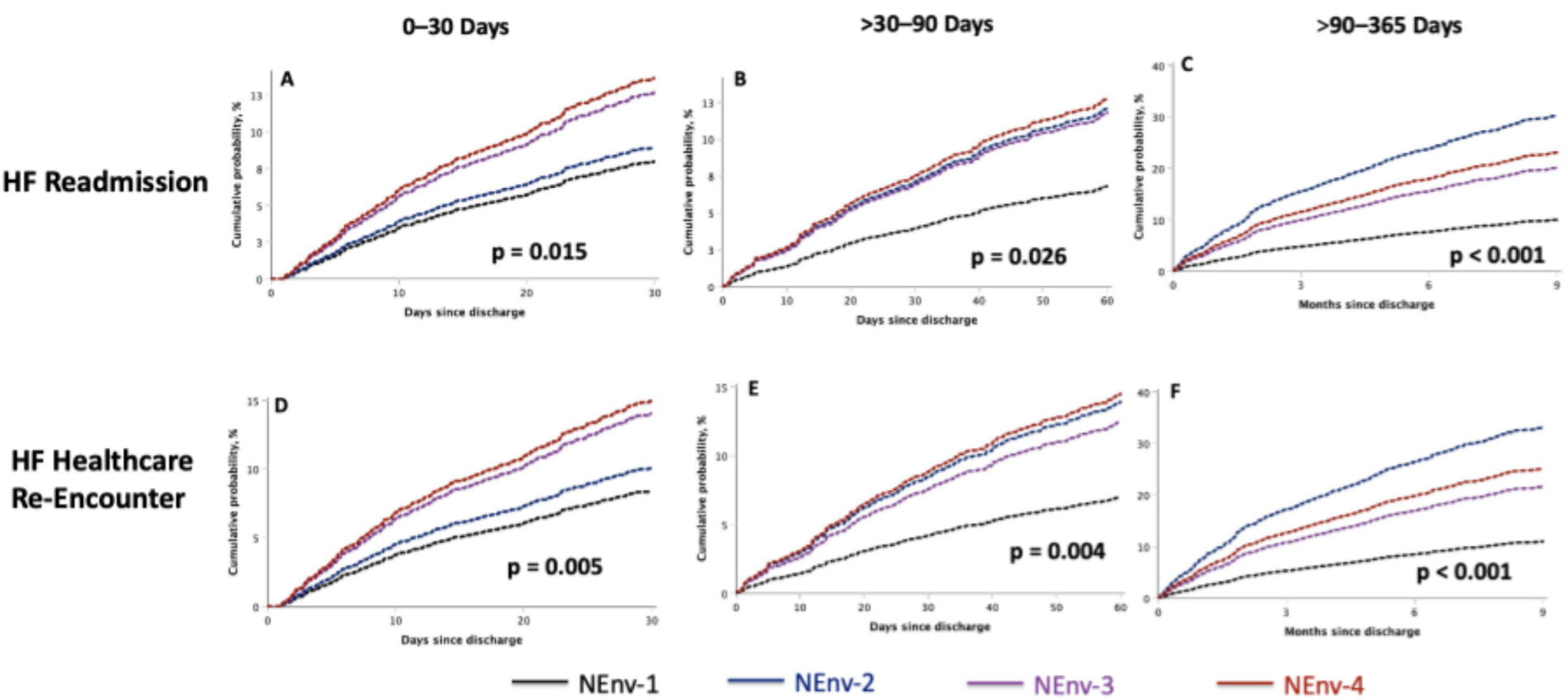
Adjusted Cumulative Probability of Heart Failure Readmission and Healthcare Re-Encounter by Neighborhood Environment. Adjusted cumulative probability curves derived from Cox proportional hazards models show time to first event following discharge from the index heart failure hospitalization. Panels A–C display heart failure readmission across prespecified post-discharge intervals (0–30 days, >30–90 days, and >90–365 days), and Panels D–F display heart failure–related healthcare re-encounters across the same intervals. Healthcare re-encounter was defined as a composite of heart failure– related emergency department visits, observation stays, or hospital readmissions. Models were adjusted for demographic and clinical covariates as described in Methods. Curves are stratified by neighborhood environment (NEnv-1 to NEnv-4), with NEnv-1 as the reference group. P values represent the overall adjusted association between neighborhood environment and each outcome within the specified interval.

**Table 2.** Association Between Neighborhood Environment and Heart Failure Readmission. Cox proportional hazards models evaluating the association between neighborhood environment (NEnv-1 to NEnv-4) and time to first heart failure readmission across prespecified post-discharge intervals (0–30 days, >30–90 days, and >90–365 days). NEnv-1 served as the reference group. Hazard ratios (HRs) are presented as unadjusted and adjusted estimates with 95% confidence intervals (CIs). Adjusted models included demographic and clinical covariates as described in Methods. **Abbreviations:** CI, confidence interval; HF, heart failure; HR, hazard ratio; NEnv, neighborhood environment.

|  | NEnv | Unadjusted HR<br>(95% CI) | P value | Adjusted HR<br>(95% CI) | P value |
| --- | --- | --- | --- | --- | --- |
| **0–30 days** |  |  |  |  |  |
|  | NEnv-1 | Reference | - | Reference | - |
|  | NEnv-2 | 1.03 (0.58–1.84) | .919 | 1.13 (0.61–2.07) | .701 |
|  | NEnv-3 | 1.59 (1.12–2.26) | .009 | 1.63 (1.12–2.37) | .011 |
|  | NEnv-4 | 1.75 (1.24–2.46) | .001 | 1.76 (1.21–2.57) | .003 |
| **>30–90 days** |  |  |  |  |  |
|  | NEnv-1 | Reference | - | Reference | - |
|  | NEnv-2 | 1.89 (1.10–3.24) | .021 | 1.83 (1.01–3.30) | .045 |
|  | NEnv-3 | 1.66 (1.11–2.46) | .013 | 1.78 (1.16–2.73) | .008 |
|  | NEnv-4 | 1.78 (1.21–2.63) | .003 | 1.94 (1.26–2.97) | .003 |
| **>90–365 days** |  |  |  |  |  |
|  | NEnv-1 | Reference | - | Reference | - |
|  | NEnv-2 | 3.46 (2.17–5.51) | <.001 | 3.42 (2.12–5.53) | <.001 |
|  | NEnv-3 | 2.32 (1.59–3.37) | <.001 | 2.13 (1.45–3.14) | <.001 |
|  | NEnv-4 | 2.80 (1.94–4.04) | <.001 | 2.49 (1.69–3.67) | <.001 |

Between 30 and 90 days after discharge, all non-reference environments were associated with increased readmission risk, including NEnv-2 (aHR, 1.83; 95% CI, 1.01–3.30; P=0.045), NEnv-3 (aHR, 1.78; 95% CI, 1.16–2.73; P=0.008), and NEnv-4 (aHR, 1.94; 95% CI, 1.26–2.97; P=0.003). Beyond 90 days, risk was highest in NEnv-2 (aHR, 3.42; 95% CI, 2.12–5.53; P<0.001), followed by NEnv-4 (aHR, 2.49; 95% CI, 1.69–3.67; P<0.001) and NEnv-3 (aHR, 2.13; 95% CI, 1.45–3.14; P<0.001).

### Heart Failure–Related Healthcare Re-Encounters

A similar pattern was observed for HF-related healthcare encounters (Figure 2D–F; Table 3). Within the first 30 days after discharge, higher risk was observed in NEnv-3 (aHR, 1.72; 95% CI, 1.20–2.47; P=0.003) and NEnv-4 (aHR, 1.84; 95% CI, 1.28–2.66; P=0.001), but not in NEnv-2.

**Table 3.** Association Between Neighborhood Environment and Heart Failure–Related Healthcare Re-Encounters. Cox proportional hazards models evaluating the association between neighborhood environment (NEnv-1 to NEnv-4) and time to first heart failure–related healthcare re-encounter across prespecified post-discharge intervals (0–30 days, >30–90 days, and >90–365 days). Healthcare re-encounters included emergency department visits, observation stays, or inpatient readmissions. NEnv-1 served as the reference group. Hazard ratios (HRs) are presented as unadjusted and adjusted estimates with 95% confidence intervals (CIs). Adjusted models included demographic and clinical covariates as described in Methods. **Abbreviations:** CI, confidence interval; HF, heart failure; HR, hazard ratio; NEnv, neighborhood environment.

|  | NEnv | Unadjusted HR<br>(95% CI) | P value | Adjusted HR<br>(95% CI) | P value |
| --- | --- | --- | --- | --- | --- |
| **0–30 days** |  |  |  |  |  |
|  | NEnv-1 | Reference | - | Reference | - |
|  | NEnv-2 | 1.18 (0.68–2.02) | .560 | 1.21 (0.68–2.16) | .520 |
|  | NEnv-3 | 1.68 (1.20–2.36) | .003 | 1.72 (1.20–2.47) | .003 |
|  | NEnv-4 | 1.88 (1.35–2.62) | <0.001 | 1.84 (1.28–2.66) | .001 |
| **>30–90 days** |  |  |  |  |  |
|  | NEnv-1 | Reference | - | Reference | - |
|  | NEnv-2 | 2.25 (1.33–3.81) | .002 | 2.06 (1.18–3.62) | .012 |
|  | NEnv-3 | 1.77 (1.19–2.64) | .005 | 1.84 (1.21–2.79) | .005 |
|  | NEnv-4 | 2.12 (1.43–3.13) | <0.001 | 2.16 (1.42–3.27) | <0.001 |
| **>90–365 days** |  |  |  |  |  |
|  | NEnv-1 | Reference | - | Reference | - |
|  | NEnv-2 | 3.63 (2.32–5.68) | <0.001 | 3.45 (2.17–5.48) | <0.001 |
|  | NEnv-3 | 2.32 (1.62–3.32) | <0.001 | 2.09 (1.44–3.03) | <0.001 |
|  | NEnv-4 | 2.81 (1.97–4.00) | <0.001 | 2.49 (1.71–3.60) | <0.001 |

Between 30 and 90 days after discharge, all non-reference environments were associated with increased risk, including NEnv-2 (aHR, 2.06; 95% CI, 1.18–3.62; P=0.012), NEnv-3 (aHR, 1.84; 95% CI, 1.21–2.79; P=0.005), and NEnv-4 (aHR, 2.16; 95% CI, 1.42–3.27; P<0.001). These associations persisted for the >90 to 365 day interval, with the greatest risk observed in NEnv-2 (aHR, 3.45; 95% CI, 2.17–5.48; P<0.001), followed by NEnv-4 (aHR, 2.49; 95% CI, 1.71–3.60; P<0.001), and NEnv-3 (aHR, 2.09; 95% CI, 1.44–3.03; P<0.001).

### Heart Failure Hospitalization Burden

The HF hospitalization burden during follow-up after the index HF hospitalization differed significantly across neighborhood environments (Figure 3; Table 4; P<.001). During follow-up, 52.3% of the patients experienced no recurrent HF admissions, 20.1% had one admission, and 27.6% had ≥2 admissions. The proportion of patients without recurrent hospitalization was the highest in NEnv-1 (72.2%) and progressively lower across NEnv-2 (53.3%), NEnv-3 (52.0%), and NEnv-4 (45.2%). Conversely, recurrent hospitalization (≥2 admissions) was the most frequent in NEnv-4 (34.1%) and least frequent in NEnv-1 (13.8%).

**Figure 3.**
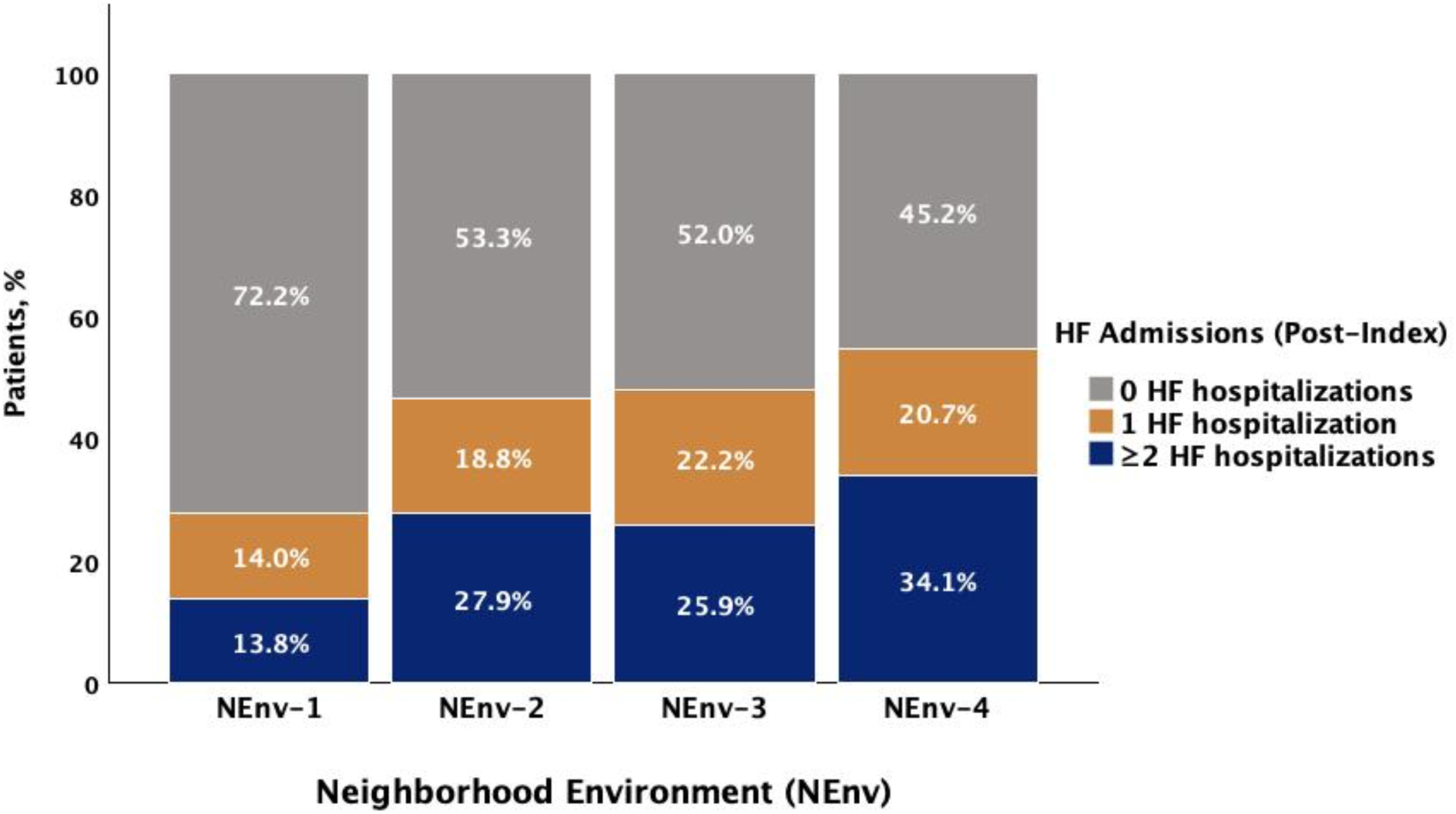
Post-Index Heart Failure Hospitalization Burden by Neighborhood Environment. Stacked bar graph showing the distribution of post-index heart failure hospitalizations (0, 1, or ≥2 admissions) across neighborhood environments (NEnv-1 to NEnv-4). Percentages are calculated within each neighborhood environment group. Group sizes were NEnv-1 (n=500), NEnv-2 (n=197), NEnv-3 (n=1191), and NEnv-4 (n=1378).

**Table 4.** Association Between Neighborhood Environment and Post-Index Heart Failure Hospitalization Burden. Multinomial logistic regression models evaluating the association between neighborhood environment and post-index heart failure hospitalization burden. Outcomes were modeled as (1) 1 versus 0 HF admissions, representing the likelihood of any readmission after the index event, and (2) ≥2 versus 1 HF admissions, representing the likelihood of recurrent hospitalization among patients with at least one readmission. NEnv-1 served as the reference group. Adjusted odds ratios (aORs) with 95% confidence intervals (CIs) are shown. Models were adjusted for covariates as described in Methods. **Abbreviations:** aOR, adjusted odds ratio; CI, confidence interval; HF, heart failure; NEnv, neighborhood environment.

| **Neighborhood environment** | **1 vs 0**<br>**HF admissions**<br>**aOR (95% CI)** | **P value** | **≥2 vs 1**<br>**HF admission**<br>**aOR (95% CI)** | **P value** |
| --- | --- | --- | --- | --- |
| NEnv-1 | Reference | - | Reference | - |
| NEnv-2 | 1.84 (1.13–3.01) | 0.015 | 1.74 (0.98–3.09) | 0.058 |
| NEnv-3 | 2.15 (1.58–2.94) | <0.001 | 1.12 (0.76–1.66) | 0.575 |
| NEnv-4 | 2.24 (1.62–3.08) | <0.001 | 1.64 (1.10–2.45) | 0.015 |

In adjusted analyses, compared with NEnv-1, all other environments were associated with higher odds of at least one post-index HF admission, including NEnv-2 (aOR, 1.84; 95% CI, 1.13–3.01; P=0.015), NEnv-3 (aOR, 2.15; 95% CI, 1.58–2.94; P<0.001), and NEnv-4 (aOR, 2.24; 95% CI, 1.62–3.08; P<0.001). Among patients with ≥1 admission, NEnv-4 was associated with higher odds of recurrent hospitalization (≥2 vs. 1 admission; aOR, 1.64; 95% CI, 1.10–2.45; P=0.015), whereas NEnv-2 showed a borderline association (aOR 1.74, 95% CI 0.98–3.09; P=.058) and NEnv-3 was not significantly associated.

## Discussion

### Principal Findings

In this cohort of patients hospitalized with HFpEF, neighborhood environment was independently associated with post-discharge HF utilization, including readmission, healthcare re-encounters, and cumulative hospitalization burden.

Two temporal patterns were observed. NEnv-3 and NEnv-4 demonstrated elevated early and sustained utilization, with higher 30-day HF readmission (aHR 1.63 and 1.76) and HF-related re-encounters (aHR 1.72 and 1.84), persisting across subsequent intervals up to >90–365 days (readmission aHR up to 2.49). In contrast, NEnv-2 exhibited a delayed-risk pattern, with the highest risk in the >90–365-day interval (readmission aHR, 3.42; re-encounter aHR, 3.45).

Neighborhood environment also differentiated the burden of hospitalization after the index admission. All non-reference environments were associated with an increased likelihood of at least one subsequent HF admission (aOR 1.84–2.24). NEnv-4 uniquely demonstrated higher odds of recurrent HF hospitalization (≥2 vs. 1 admission; aOR, 1.64), indicating greater recurrence of events.

These utilization patterns corresponded with the differences in clinical profiles. NEnv-4 had the highest burden of cardiometabolic comorbidities, including diabetes mellitus (34.8%), chronic kidney disease (28.2%), obesity (44.8%), high systolic blood pressure (136.5 mm Hg), and low eGFR (47.6 mL/min/1.73m²). In contrast, NEnv-1, characterized by older age but a lower cardiometabolic burden (diabetes mellitus, 23.0%; chronic kidney disease, 19.1%), demonstrated the lowest post-index HF utilization. Overall, the neighborhood environment stratifies both the timing and intensity of post-discharge HF utilization.

### Prior Literature Review

Neighborhood disadvantages have been consistently associated with increased HF readmission and mortality, particularly during the early post-discharge period, with disparities amplified across racial and socioeconomic groups.^15,16,21^ This early excess risk reflects the vulnerable phase following HF hospitalization, during which residual congestion, clinical instability, and incomplete care transitions contribute to recurrent events.^6,7^ The present results build on this framework by showing that post-discharge risk varies across neighborhood environments and follows unique temporal patterns.

Whereas prior studies have emphasized early readmission risk,^6,15,16^ we identified a delayed risk trajectory in NEnv-2. Despite sharing rural characteristics with NEnv-1, NEnv-2 exhibited the highest late risk of HF readmission and healthcare re-encounter at 1 year. This divergence indicates that rurality alone does not account for risk patterns and that differences in access, care continuity, and chronic disease infrastructure likely contribute.^17^ Early events appear to reflect post-discharge instability,^6,7^ whereas events in the >90–365-day interval likely reflect cumulative deficits in longitudinal disease management, including limited access to specialty care and fragmented follow-up.^7,8,17^

The persistent and recurrent utilization of NEnv-4 is consistent with a greater burden of cardiometabolic disease and multimorbidity. Diabetes mellitus, chronic kidney disease, obesity, hypertension, and reduced kidney function are established drivers of recurrent HF hospitalization and sustained healthcare utilization.^9^ The co-occurrence of cardiometabolic comorbidity and adverse social conditions likely contributes to ongoing vulnerability beyond the immediate post-discharge period.

The lower observed utilization of NEnv-1 should be interpreted in the context of competing risks. Prior work in this cohort demonstrated higher mortality in NEnv-1,^20^ suggesting that earlier death may limit the opportunity for subsequent hospitalization. This is consistent with HF literature demonstrating that mortality and rehospitalization are interrelated but competing outcomes.^22^

Across environments, HF utilization reflected both the likelihood of initial readmission and, in NEnv-4, a greater propensity for recurrent hospitalization. These findings highlight the importance of evaluating total hospitalization burden and how risk evolves over time, rather than relying solely on time-to-first-event metrics.^9^

### Clinical Impact

The timing of excess HF risk differed across neighborhood environments, suggesting that a single post-discharge strategy may not adequately address the needs of all patient groups. Patients in NEnv-4 and, to a lesser extent, NEnv-3 experienced higher early utilization, which suggests that earlier follow-up, support with medication access, and closer outpatient coordination may be beneficial in these groups.^2,7^ In contrast, the later increase in risk observed in NEnv-2 suggests a need for extended follow-up, with emphasis on improved access to specialty care, telehealth-supported follow-up, and ongoing disease monitoring.^8,9^

The interpretation of utilization patterns also requires the consideration of competing outcomes. The lower observed hospitalization rates in the NEnv-1 group may, in part, reflect higher mortality, as previously demonstrated in this cohort. ^20^ This underscores the importance of evaluating HF outcomes in the context of survival. More broadly, the observed differences across neighborhood environments suggest that neighborhood context may be useful for risk stratification and for tailoring interventions to how HF utilization changes over time.

### Strengths and Limitations

This study has several strengths. It includes a large contemporary cohort of patients with HFpEF with detailed clinical characterization and evaluates multiple complementary measures of HF utilization across clinically relevant post-discharge intervals (0–30, >30–90, and >90–365 days). The use of a machine-learning-based neighborhood framework capturing multidimensional social determinants enabled the identification of distinct patterns of post-discharge risk that were not apparent with single-domain measures. The consistency of findings across outcomes and intervals supports the robustness of the results.

This study has some limitations. The observational retrospective design introduced the potential for residual confounding factors. Data on key factors related to healthcare access and post-discharge care, including insurance status, outpatient follow-up, and medication adherence, were unavailable. Neighborhood-level ZIP code assignment may not reflect individual-level exposure and may introduce potential ecological misclassifications. A single-center design may limit generalizability. In addition, mortality was not modeled as a competing risk, which may have influenced the interpretation of utilization patterns, particularly in NEnv-1.

### Conclusion

Neighborhood environment was associated with variation in the timing of post-discharge HF utilization in HFpEF, including early risk (0–30 days) in NEnv-3 and NEnv-4 and later risk (>90–365 days) in NEnv-2, as well as differences in recurrent hospitalization. These results indicate that incorporating neighborhood context into risk assessment may help identify when patients are most vulnerable after discharge and guide the timing of post-discharge care.

## Author Contribution

**YA:** Conceptualization, methodology, formal analysis, software, visualization, writing – original draft, writing – review, and editing. **ER:** Data curation, writing, reviewing, and editing. **CS:** Writing, review, and editing. **JTB:** Methodology, writing, review, and editing. **JEL:** Methodology, Writing, Review, and Editing. **MN:** Methodology, formal analysis, writing, review, and editing. **IE:** Methodology, writing, reviewing, and editing. **LI:** Methodology, writing, review, and editing. **DAL:** Methodology, writing, review, and editing. **NC:** Conceptualization, methodology, supervision, resources, funding acquisition, writing, reviewing, and editing. **MC:** Conceptualization, methodology, supervision, project administration, resources, funding acquisition, writing, review, and editing. All authors have contributed to the manuscript and approved the submitted version.

## Acknowledgments

We thank the patients and staff of UC Davis Health for their contributions to this research. We acknowledge the use of publicly available data from the American Community Survey.

## Data Availability Statement

The data underlying this study cannot be shared publicly because of privacy concerns regarding patient health information. American Community Survey data were obtained from the US Census Bureau. Requests to access aggregated and de-identified data for academic purposes may be submitted to the corresponding author and approved in accordance with institutional policies.

## Non-standard Abbreviations and Acronyms

ACEi: angiotensin-converting enzyme inhibitor
ACS: American Community Survey
ADI: Area Deprivation Index
AF/AFL: atrial fibrillation or atrial flutter
ARB: angiotensin receptor blocker
BMI: body mass index
BNP: brain natriuretic peptide
CKD: chronic kidney disease
COPD: chronic obstructive pulmonary disease
HF: heart failure
HFpEF: heart failure with preserved ejection fraction
HR: hazard ratio
ICD: International Classification of Diseases
IHD: ischemic heart disease
IQR: interquartile range
LVEF: left ventricular ejection fraction
MRA: mineralocorticoid receptor antagonist
NEnv: neighborhood environment
SDoH: social determinants of health
SGLT2i: sodium-glucose cotransporter 2 inhibitor
ZCTA: ZIP Code Tabulation Area

